# The heart of an endurance athlete: impact of and recovery after an ultra-endurance event

**DOI:** 10.1101/2020.07.16.20155127

**Authors:** Anton Swart, Demitri Constantinou

## Abstract

**Aims:** Acute bouts of ultra-endurance exercise may cause an acute reduction in cardiac function, causing a physiological cascade which releases cardiac biomarkers. This study set out to determine the cardiac stress and recovery of participation in a three-day ultra-endurance mountain biking event of athletes using heart rate variability (HRV) as an outcome measure.

**Methods:** Sixteen healthy participants (male and female) participating in a three-day ultra-endurance mountain biking event underwent a five-minute resting ECG recording in a supine position. Heart rate variability measurements were recorded two days before the race (baseline testing), after each race day, and at 24-hour post-event (recovery).

**Results:** Time-domain and frequency domain measures showed significant (p≤0.05) changes from baseline in HRV parameters after each race day. The significant changes in HRV parameters reflected an increase in sympathetic activity after each day of the event. Our data revealed that the mean HR and RR variability variables did not return to baseline value after 24-hours of recovery, reflecting autonomic nervous system dysfunction, and that changes persisted for at least 24-hours post-event.

**Conclusion:** Our study shows that competing in an ultra-endurance mountain bike event led to diminished vagal activity and a decrease in HRV throughout the event and persisted for at least 24-hours post-event. The body was under continuous sympathetic dominance during rest as well as during each day of racing, implying each race day can be considered a physiological stress. This may, in turn, cause a disturbance in homeostasis and an increase in autonomic nervous system dysfunction. This has implications for further research, including dysrhythmia risk, and monitoring of athletes in advising a return to strenuous activity.

## 1. Introduction

There is every belief, substantiated by research, that exercise is good for one. What is unknown is whether there is a significant impact on the heart of an endurance athlete after completing an ultra-endurance mountain bike event.

The evidence is that the impact of exercise leads to healthy physiological adaptations and that most endurance athletes benefit from it.^1^ The healthy heart of an endurance athlete can respond effectively to acute exercise and can delay fatigue during prolonged exercise.^1^ George et al (2012) studied the acute and chronic adaptation on endurance athletes’ hearts and shown that acute bouts of ultra-endurance exercise may cause an acute reduction in cardiac function, causing a physiological cascade which releases cardiac biomarkers. Thus, this evidence indicates that some endurance athletes may develop a pathophysiological cascade. Athletes and clinicians should be mindful of it and react adequately to this pathophysiological phenomenon.^1^

Heart rate variability (HRV) monitoring is a useful indicator in the diagnosis and prevention strategies of over-reaching in athletes.^2^ Meeusen et al (2013) defined over-reaching as an “accumulation of training and/or non-training stress resulting in a short-term decrement in performance capacity with or without related physical and psychological signs and symptoms of maladaptation in which restoration of performance capacity may take from several days to several weeks”.

Often over-reaching is related to several warning signs, one of which includes autonomic nervous system (ANS) dysfunction and imbalances.^3^ It is therefore known that the physiological system can be compromised due to an increase in exercise stress either from increased intensity and/or from the duration of exertion.^4^ Thus the purpose of this study was to determine the effect of participation in a three-day ultra-endurance mountain bike event on the heart of athletes, using the outcome measure of HRV as a marker of autonomic function. The hypothesis was that there would be significant variations in HRV during and/or after the three days of competing in an ultra-endurance mountain biking event and HRV would not fully recover to baseline values within 24-hours.

## 2. Methods

### 2.1. Study Design

The study used a prospective quantitative research design, which involved the collection and analysis of numerical data. All data underlying this article will be shared on reasonable request to the corresponding author. None sources of funds were received by a second party and there is no competing of interest between authors and is none declared.

For the duration of retention of records, each participant’s health records and measurements are held by following the guidelines of the Health Professional Council of South Africa (HPCSA) and Health Professional Council of Namibia (HPCNA). All hard copy records are stored in a safe place with electronic copies uploaded to online cloud storage, safeguarded by a password. All records and measurements are stored for a period of not less than six years and only the principle researcher has access to these records. Indirect identifiers (numbers) replaced all obvious identifiers (example name and surname) of participants in the main data set to retain confidentiality. A separate data set known as the “Key” holds the indirect identifiers with the obvious identifiers.

### 2.2. Study population and process

The study comprised of a sample of 17 participants (male and female) who partook in a three-day ultra-endurance mountain biking event and were selected through a sample of convenience. The race organizers of the Standard Bank Klein-Aus Vista Mountain Bike Challenge approved and provided written permission to conduct the study during their event.

One participant failed to complete the event on the third day and was excluded from the study. Thus, data were collected on 16 participants that completed the full three-day stage event. This study confirms with the principles outlines in the Declaration of Helsinki.

Participants had the opportunity to volunteer to take part in the study after race organizers sent an invitation via email. The researcher thereafter arranged a meeting and presented to the participants details about the study. During the meeting, an informed consent form was available and participants had the opportunity to read, understand and sign the consent form before being enrolled in the study. Ethical considerations were observed and ethics approval was granted by the Biomedical Research Ethics Committee (BREC) and Research Management Committee (RMC), Ministry of Health and Social Services in Namibia and the Humans Research Ethics Committee (Medical) of the University of the Witwatersrand, South Africa (clearance certificate M171037).

All participants were encouraged to avoid drinking alcoholic beverages and avoid smoking before and during the time of the study. No participant reported taking any medication or sympathomimetic drugs that could have affected the cardiovascular system. All participants were athletic, healthy and amateur mountain bike riders with marathon mountain bike riding experience of at least three years.

### 2.3. Characteristics of the three-day mountain bike race

The event consisted of a three-day mountain bike stage event covering a total distance of ∼140km. This event, set in Namibia, is highly technical, and only advised for highly experienced riders. The event took place during high temperatures of 37-40 degrees Celsius, with a total elevation gain of +-3100m across the three days shown in Figure 1. The average finishing time was nine hours.

**Figure 1.**
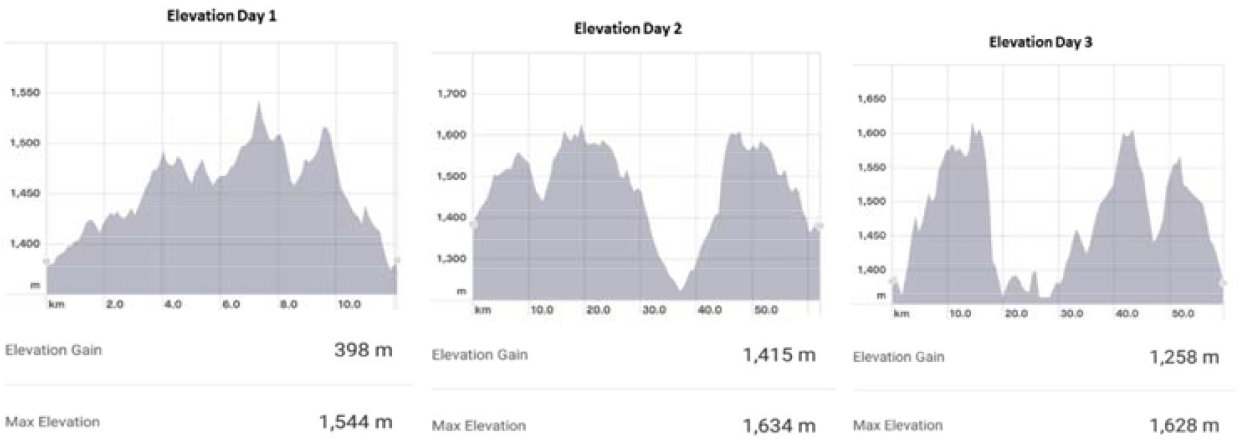
Elevation gain and maximal elevation of day one, two and three.

### 2.4. Heart Rate Variability Variables

The simplest and most valid method to evaluate HRV is through the measurement of time-domain measures during which the intervals between successive normal complexes are determined.^4,8,9^ Frequency-domain consists of various spectral methods to calculate and estimate the R-R intervals in series, which calculates the distribution of absolute and relative power into different frequency bands. This provides simple data of how power distributes as a function of frequency.^4,8,9^ A summary of time domain and frequency domain measures are presented in Table 1.

**Table 1.** HRV variables

| **Time domain measures:** | **Frequency domain measures:** |
| --- | --- |
| Mean Heart Rate | Total Power (TP) of overall RR-variability |
| Mean RR-variability | High-Frequency Power (HF) (0.15-0.40Hz) |
| Standard deviation of the sequence of NN intervals (SDNN) | Low-Frequency Power (LF) (0.04-0.15Hz) |
| Number of pairs of successive RR interval which differ more than 50 ms (NN50) | Very Low-Frequency Power (VLF) ($\leq 0.04$Hz) |
| Square root of the mean squared differences between successive RR intervals (RMSSD) | Low-Frequency power (LF)/High-Frequency Power (HF), the ratio between LF and HF powers |

### 2.5. HRV Analysis

Welch Allyn (Oregon, USA) PC based stress Electrocardiography (ECG) system was used to obtain a resting 12 lead standard ECG tracing. The HRV module in the Cardio Perfect software (version 1.6.6.1146) analysed short-term (five-minute recording) HRV in a series of heartbeat intervals up to a maximal resting ECG recording of five minutes and is validated software for R-R-variability analysis.^5^ This module automatically detected the QRS complexes from which the R-R intervals are constructed. The R-R intervals were displayed along with the ECG for assessment of QRS classifications and localization of measurement artefacts and noise.

### 2.6. Data Collection

#### 2.6.1. Baseline testing procedures

Two days before the event, body composition of all participants was measured making using of the International Society for the Advancement of Kinanthropometry (ISAK) standards.^6^ Participants underwent a pre-ECG reading stabilization period of rest for five minutes to ensure that heart rate levelled before recording HRV. This was followed with a resting ECG recording in a supine position for five-minutes which provided baseline (pre-event) values. Electrode placement was done by following the standard 12 lead ECG electrode placement method.^7^ During the five-minute recording, the athlete lay still to decrease interference and noise. An overview of the study’s data collection from baseline to 24-hour post-event measure is shown in Figure 2.

**Figure 2.**
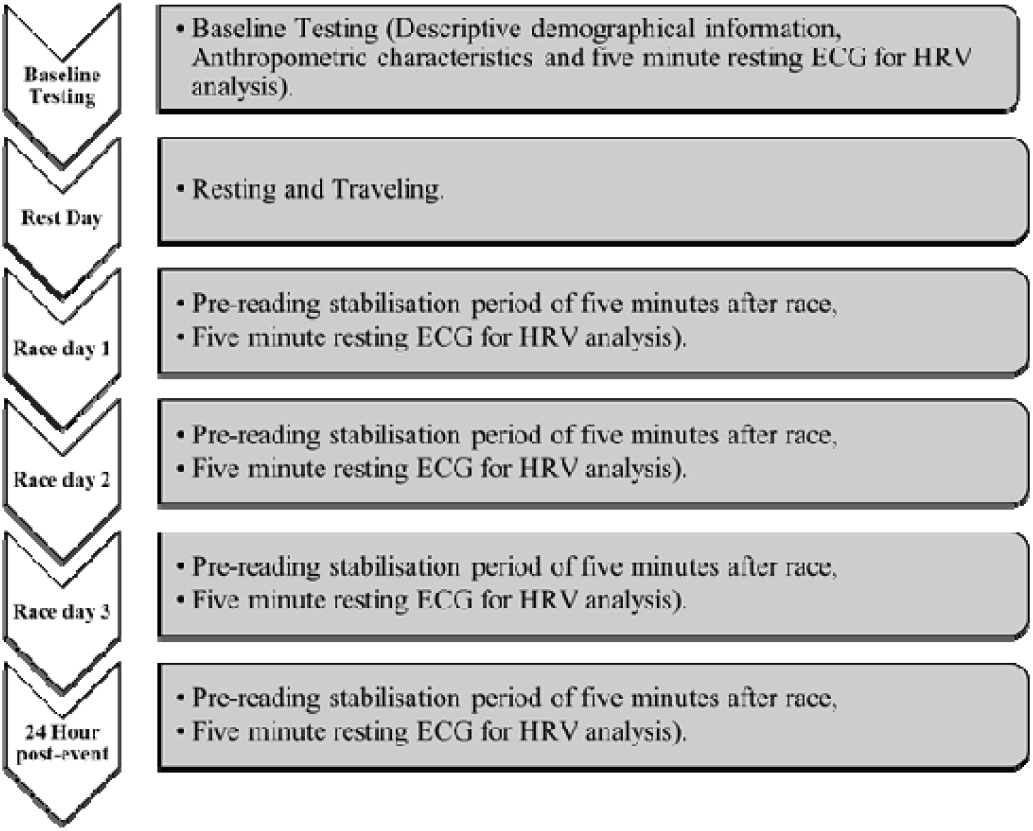
Overview of the study’s data collection.

#### 2.6.2. Daily procedures

Participants presented to a designated area once crossing the finishing line after each stage for the post-event measurement procedures. They presented within two-minutes after crossing the finishing line and underwent a pre-reading stabilization period for five-minute before recording HRV, through means of a five-minute resting ECG in a supine position.

#### 2.6.3. 24-hour Post-event testing procedures

A 24-hour post-event measurement was recorded at Klein-Aus Vista the day after the participants had completed the third and final stage of the event. Participants underwent a pre-reading stabilization period of five-minute rest before recording an ECG.

### 2.7. Statistical analysis

The descriptive data are expressed and summarized as means ± SD and were analysed using the statistical package STATISTICA (version 13.2). The normality of HRV distribution was checked with the Shapiro-Wilk normality test, which confirmed non-normality. The study used non-para-metric tests, which included Friedman two-way analysis of variance and Wilcoxon signed-rank test. Results are expressed as median and interquartile range. Statistical significant was set at p ≤ 0.05.

## 3. Results

The descriptive demographic information and anthropometric characteristics of the 16 participants are summarized in Table 2. Before data processing all ECG tracing measurements were automatically corrected for artefact removal through the Welch Allyn Cardio Perfect software for ectopic and missed beats, obtaining the normal-to-normal interval (NN).

**Table 2.** Descriptive demographical information and anthropometric characteristics of participants (n=16).

| **Variable** | **Mean ± SD (range)** |
| --- | --- |
| **Age (years)<sup>a</sup>** | 48.75 ± 7.41 (39-60) |
| **Body Weight (kg)<sup>a</sup>** | 78.48 ± 12.18 (63-100) |
| **Height (m)<sup>a</sup>** | 174.31 ± 6.90 (166-187) |
| **BMI (m/kg<sup>2</sup>)<sup>a</sup>** | 25.71 ± 2.64 (22-30) |
| **Body Fat %<sup>a</sup>** | 12.94 ± 3.28 (7-18) |
| **Waist: hip ratio<sup>a</sup>** | 0.86 ± 0.06 (0.74-0.92) |

<sup>a</sup>Data are presented as mean ± SD. Abbreviation: BMI = body mass index

### 3.1. Time domain measures

Compared with baseline testing, mean heart rate (HR) increased and R-R variability decreased significantly (Table 3) and continued to increase and decrease until after day two and three of the event respectively, but did not recover back to baseline measurements 24-hour post-event. During the 24-hour post-event, mean HR and R-R-variability measured significantly higher and lower respectively compared to baseline measurements (Table 3). Box and Whisker Plot of HR and R-R variability indicative of minimum, quartile one, median, quartile three and maximum are shown in Figure 3 and Figure 4, reflecting significance from baseline through to 24-hour post-event.

**Table 3.**
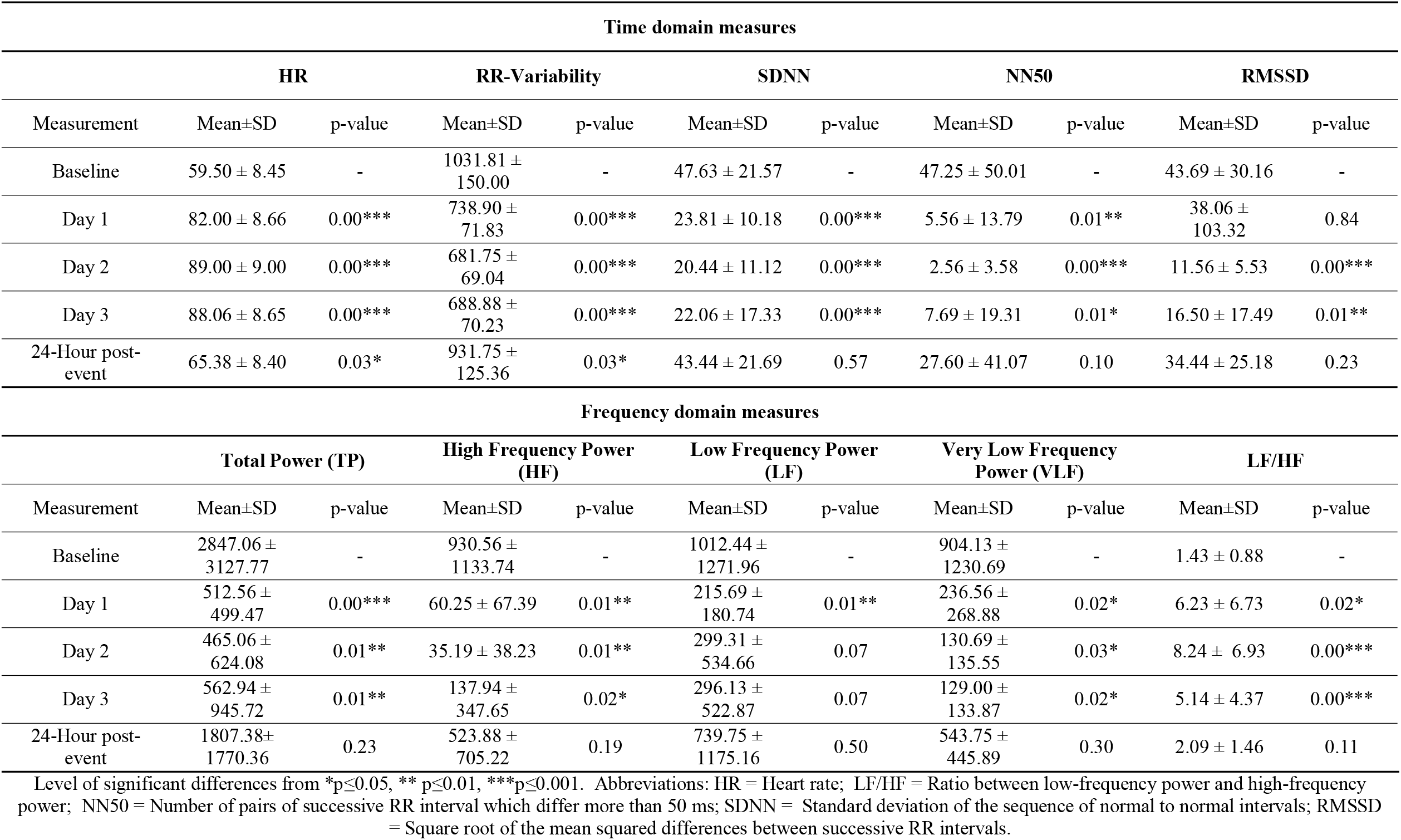
Changes in Time domain and Frequency domain measures from baseline to 24-hour post-event (n=16).

| Time domain measures |  |  |  |  |  |  |  |  |  |  |
| --- | --- | --- | --- | --- | --- | --- | --- | --- | --- | --- |
| Measurement | HR |  | RR-Variability |  | SDNN |  | NN50 |  | RMSSD |  |
|  | Mean±SD | p-value | Mean±SD | p-value | Mean±SD | p-value | Mean±SD | p-value | Mean±SD | p-value |
| Baseline | 59.50 ± 8.45 | - | 1031.81 ± 150.00 | - | 47.63 ± 21.57 | - | 47.25 ± 50.01 | - | 43.69 ± 30.16 | - |
| Day 1 | 82.00 ± 8.66 | 0.00*** | 738.90 ± 71.83 | 0.00*** | 23.81 ± 10.18 | 0.00*** | 5.56 ± 13.79 | 0.01** | 38.06 ± 103.32 | 0.84 |
| Day 2 | 89.00 ± 9.00 | 0.00*** | 681.75 ± 69.04 | 0.00*** | 20.44 ± 11.12 | 0.00*** | 2.56 ± 3.58 | 0.00*** | 11.56 ± 5.53 | 0.00*** |
| Day 3 | 88.06 ± 8.65 | 0.00*** | 688.88 ± 70.23 | 0.00*** | 22.06 ± 17.33 | 0.00*** | 7.69 ± 19.31 | 0.01* | 16.50 ± 17.49 | 0.01** |
| 24-Hour post-event | 65.38 ± 8.40 | 0.03* | 931.75 ± 125.36 | 0.03* | 43.44 ± 21.69 | 0.57 | 27.60 ± 41.07 | 0.10 | 34.44 ± 25.18 | 0.23 |
| Frequency domain measures |  |  |  |  |  |  |  |  |  |  |
| Measurement | Total Power (TP) |  | High Frequency Power (HF) |  | Low Frequency Power (LF) |  | Very Low Frequency Power (VLF) |  | LF/HF |  |
|  | Mean±SD | p-value | Mean±SD | p-value | Mean±SD | p-value | Mean±SD | p-value | Mean±SD | p-value |
| Baseline | 2847.06 ± 3127.77 | - | 930.56 ± 1133.74 | - | 1012.44 ± 1271.96 | - | 904.13 ± 1230.69 | - | 1.43 ± 0.88 | - |
| Day 1 | 512.56 ± 499.47 | 0.00*** | 60.25 ± 67.39 | 0.01** | 215.69 ± 180.74 | 0.01** | 236.56 ± 268.88 | 0.02* | 6.23 ± 6.73 | 0.02* |
| Day 2 | 465.06 ± 624.08 | 0.01** | 35.19 ± 38.23 | 0.01** | 299.31 ± 534.66 | 0.07 | 130.69 ± 135.55 | 0.03* | 8.24 ± 6.93 | 0.00*** |
| Day 3 | 562.94 ± 945.72 | 0.01** | 137.94 ± 347.65 | 0.02* | 296.13 ± 522.87 | 0.07 | 129.00 ± 133.87 | 0.02* | 5.14 ± 4.37 | 0.00*** |
| 24-Hour post-event | 1807.38 ± 1770.36 | 0.23 | 523.88 ± 705.22 | 0.19 | 739.75 ± 1175.16 | 0.50 | 543.75 ± 445.89 | 0.30 | 2.09 ± 1.46 | 0.11 |

Level of significant differences from \*p<0.05, \*\* p<0.01, \*\*\*p<0.001. Abbreviations: HR = Heart rate; LF/HF = Ratio between low-frequency power and high-frequency power; NN50 = Number of pairs of successive RR interval which differ more than 50 ms; SDNN = Standard deviation of the sequence of normal to normal intervals; RMSSD = Square root of the mean squared differences between successive RR intervals.

**Figure 3.**
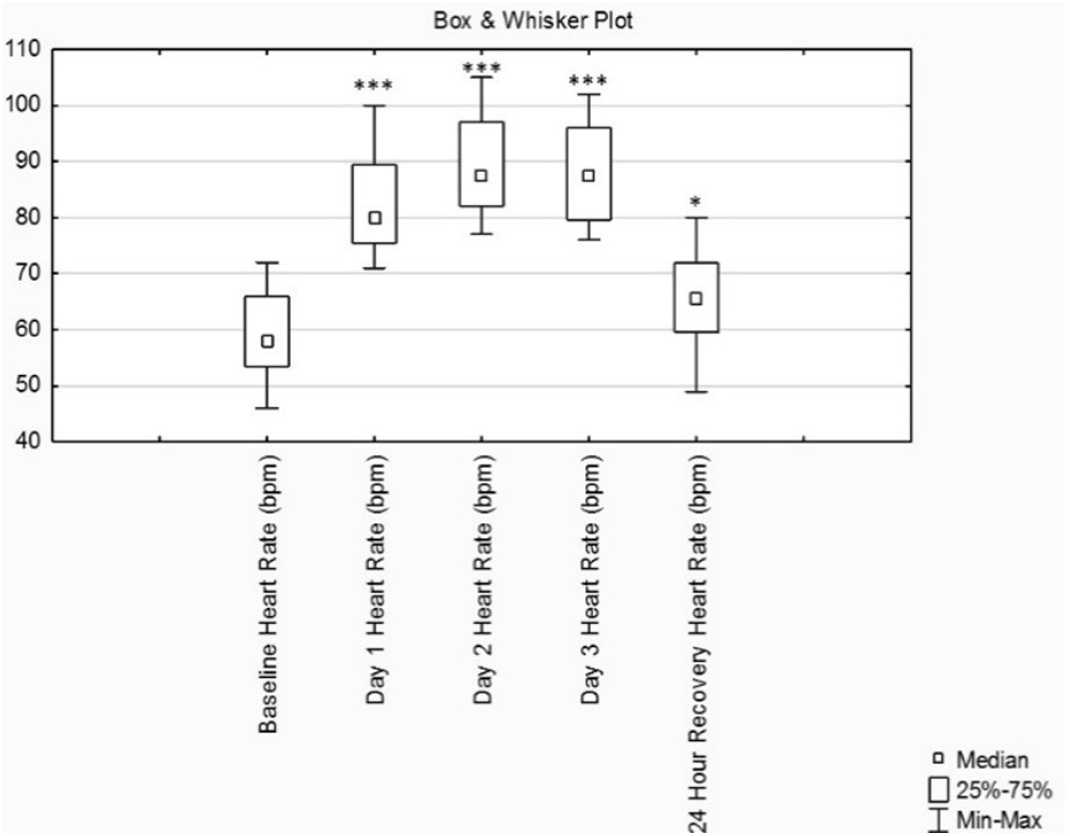
Box & Whisker Plot of the Heart Rate (n=16). Significant differences *p≤0.05, ** p≤0.01, ***p≤0.001.

**Figure 4.**
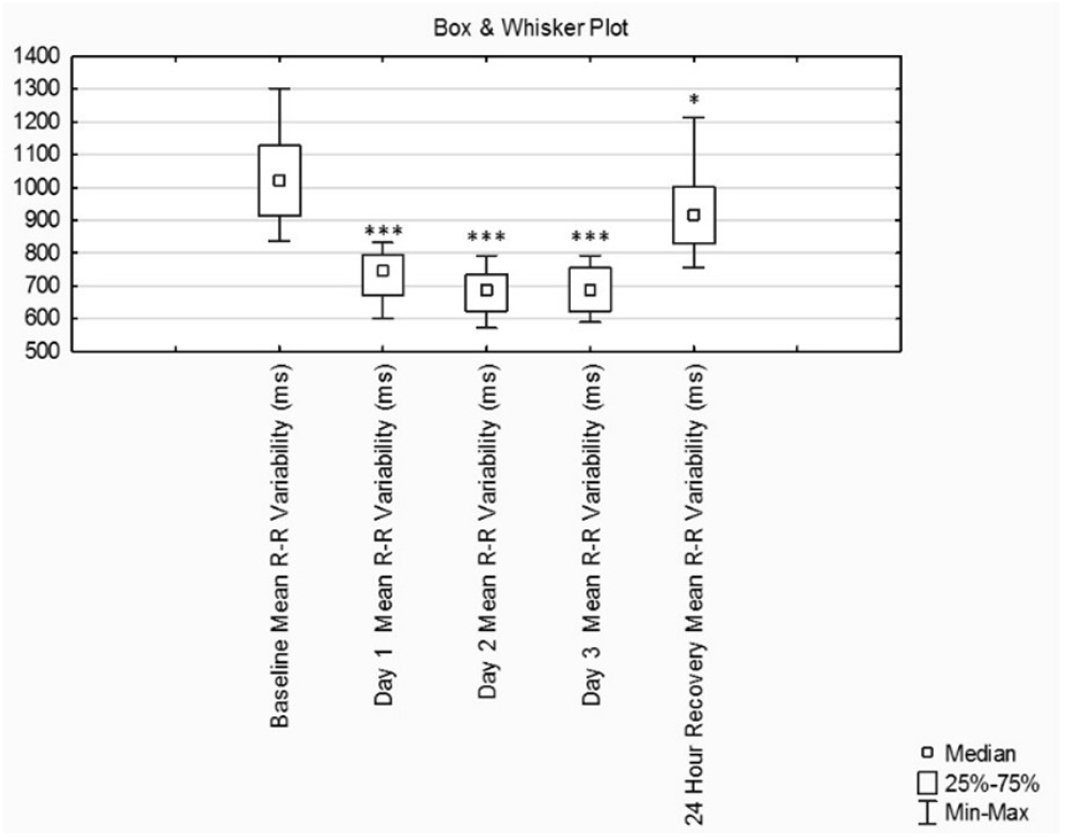
Box & Whisker Plot of the Mean R-R Variability of HRV (n=16). Significant differences *p≤0.05, ** p≤0.01, ***p≤0.001.

The interval measures of both SDNN and NN50 decreased significantly from day one compared to baseline (Table 3) and continued to decrease until after day two. After day three, there was a slight increase compared to day two but significantly lower compared to baseline values (Table 3). The 24-hour recovery revealed no significant difference between baseline and 24-hour post-event. The RMSSD decreased significantly (Table 3) on day two compared to baseline and continued to decrease until after day three. After day three, there was a slight increase in power compared to day two but significantly lower compared to baseline values (Table 3). The 24-hour post-event RMSSD value revealed no significant difference between baseline and 24-hour post-event.

### 3.2. Frequency domain measures

Changes in TP, HF, VLF and LF/HF indicative of cardiovascular modulation were associated with a significant (Table 3) change in HRV from day one compared to baseline and continued to change significantly until after day three. There were no statistically significant differences between baseline and 24-hour post-event. The LF decreased significantly (Table 3) from baseline after day one, but there were no further changes measured after days two and three, and at 24-hour post-event.

## 4. Discussion

Little research exists in this field of the recovery dynamics of HRV after endurance exercise.^10^ In recent studies by Kaikkonen, Nummela, and Rusko (2007) and Perkins et al (2017), the key factors determining post-exercise HRV appeared to be the exercise intensity and exercise duration.

### 4.1. Effects on Heart Rate response after an ultra-endurance event

In our study, the HR at post-event measures at day one, day two, day three and 24-hour post-event remained higher compared to that of the baseline values. This resultant increased sympathetic activity increases electrical activity and stress on the heart through the sympathetic nerves. Our findings suggest that after an ultra-endurance event there is some form of continued sympathetic stimulation and that the sympathetic drive continues well into the recovery phase. Thus it can be deduced that there is increased cardiac contractility at rest as from after day one of the endurance event, and up to at least 24-hours post-event.

### 4.2. Effects on Time domain measures

Time domain measures showed decreased vagal tone and increase in sympathetic tone at rest after each race day which significantly changed (decreased) up to the final day. Two other studies reported a decrease in RR intervals, SDNN, NN50 and RMSSD during endurance events, which was indicative of an increase in sympathetic activity and decreased parasympathetic activity^12,13^ – in line with the findings of our study.

### 4.3. Effects on Frequency domain measures

Our results further showed a significant decrease in TP over the three days of the event. As previously noted the resting HR post-event remained higher compared to that of the baseline values. Due to the increase in work load, physiological stress on the body increased the sympathetic drive, leading to the increased resting heart rate. Thus, the resultant relative tachycardia resulted in a reduction in TP, leading to a significant decrease in HRV across the three days of the event.

Low VLF power not only predicts autonomic dysfunction but is also indicative of an increase in inflammation.^14^ This increase in inflammation is often seen due to sympathetic response, and promotes repairing of exercise-induced myocardial and skeletal muscle damage.^14^ In our study one notices the change after day two, where there is a small change from sympathetic dominance towards parasympathetic dominance. The sympathetic system increases metabolic function to cope with exogenous challenges, while the parasympathetic (vagal) system will start to dominate to increase functions associated with growth and repair in the body.^14^

Therefore, although through the study period there was a decrease in power and the power started to plateau, one may link this to the increase in inflammation reflected in the VLF power. This may indicate inflammatory-modulated promotion of healing and repair of exercise-induced myocardial and musculoskeletal damage as from day two and continues through day three.

The LF/HF ratios from a five-minute ECG recording in a supine position measuring greater than 2.0 is an expression of the sympatho-vagal balance and prevalence of increased sympathetic activity.^15^ Our study showed a disturbance and significant increased sympathetic activity as reflected by the LF/HF ratio after day one, day two and day three. This may result in a decrease in performance and an increase in inflammation, indicated by ANS dysfunction. Therefore adequate recovery is necessary to promote repair and healing and maintain/improve performance. Our study showed that the greatest disturbance occurred after day two (8,24±6,93). The ANS began to improve after day three (5,15±4,37) but remained blunted. This is indicative that although the participants were at rest, their ANS maintained an increase in sympathetic drive.

The HF component is mediated by the parasympathetic nervous system.^8^ A decrease in HF power in our study is indicative of decreased vagal activity, reflecting an increase in an undesirable stimulus due to the extreme event conditions and the duration and intensity of the ultra-endurance event.

### 4.4. Recovery time and HRV

Both Hautala et al (2001) and Dong (2016) have addressed the implication that may arise during persistent abnormal changes in autonomic regulation: that there is an increased risk in sudden cardiac death in athletes who present with a diminished vagal activity, indicated by a decrease in HRV with the variability becoming constant.^16^ Thus recovery to baseline is prudently important.

### 4.5. Limitations

The main limitation of this study would be the assessment up to 24-hours post-event and not beyond; which in our study showed that the vagal activity remained blunted up to at least 24-hours after recovery. Bernardi et al (1996); Hautala et al (2001) and Gratze et al (2005) showed that cardiac parasympathetic activity returned to baseline levels after 24-hours of recovery and that complete cardiac autonomic recovery can take up to 48-hours of recovery.

Our study provides the following information regarding HRV and a three-day bike ultra-endurance event:

- The data provided supports the notion that 24-hours is not adequate for the full recovery of the ANS during a multi-stage and after an ultra-endurance event. Therefore, it would be wise when assessing recovery after a single ultra-endurance event that one should assess recovery beyond 48-hours. This needs further research.
- That HRV can be used to analyse the stress of the ultra-endurance event on the ANS, which in our study showed to be significant after day one and continued through days two and three of the event.
- The ANS and dysfunction correlating to sudden cardiac arrest suggests that healthcare and emergency personnel should be particularly aware of a possible increased risk on day two of an ultra-endurance event.

Therefore, we recommend making use of HRV as a measuring tool to assess cardiac autonomic activity, during which sports physicians, athletes, and coaches can assess the stress of ultra-endurance events on the ANS and plan for correct recovery strategies after a single ultra-endurance event. Furthermore, we recommend that HRV should not only be used to asses recovery but should be used as a screening tool where sports physicians can assess the state of the ANS before an ultra-endurance event. The reasoning is that should the ANS be in a state of dysfunction, susceptible athletes might be at risk for a cardiovascular event. Further research is needed to determine how HRV measurements may be used in reducing cardiac events in individuals with an increased risk.^17^

## Supporting information

STROBE Checklist

## Data Availability

All data underlying this article will be shared on reasonable request to the corresponding author. For the duration of retention of records, each participants health records and measurements are held by following the guidelines of the Health Professional Council of South Africa (HPCSA) and Health Professional Council of Namibia (HPCNA). All hard copy records are stored in a safe place with electronic copies uploaded to online cloud storage, safeguarded by a password. All records and measurements are stored for a period of not less than six years and only the principle researcher has access to these records. Indirect identifiers (numbers) replaced all obvious identifiers (example name and surname) of participants in the main data set to retain confidentiality. A separate data set known as the Key holds the indirect identifiers with the obvious identifiers.

## 6. Acknowledgments

This study was undertaken at the University of the Witwatersrand, South Africa. The principal author thank the following for their assistance and contribution to the development and achievement of this research:

Mr. A. da Silva a technical service engineer from Africa Service & Solutions with assistance in the Welch Allyn Cardio-Perfect software, which was used during the data collection for this study;

P. Swiegers (Chief Race Official and Organizer) at Klein-Aus-Vista lodge, Namibia, to make use of his event to conduct this research study.

## 8. Authors’ contributions

A.S. participated in the design of the study, contributed to data collection, data analysis and interpretation of results, and contributed to the writing of the manuscript; D.C. participated in the writing of the manuscript and supportive feedback and supervision during the research study. All authors have read and approved the final version of the manuscript, and agree with the order of presentation of the authors.

